# Generation and transmission of inter-lineage recombinants in the SARS-CoV-2 pandemic

**DOI:** 10.1101/2021.06.18.21258689

**Authors:** Ben Jackson, Maciej F Boni, Matthew J Bull, Amy Colleran, Rachel M Colquhoun, Alistair Darby, Sam Haldenby, Verity Hill, Anita Lucaci, John T McCrone, Samuel Nicholls, Áine O’Toole, Nicole Pacchiarini, Radoslaw Poplawski, Emily Scher, Flora Todd, Hermione Webster, Mark Whitehead, Claudia Wierzbicki, The COVID-19 Genomics UK (COG-UK) consortium, Nicholas J Loman, Thomas R Connor, David L Robertson, Oliver G Pybus, Andrew Rambaut

## Abstract

We present evidence for multiple independent origins of recombinant SARS-CoV-2 viruses sampled from late 2020 and early 2021 in the United Kingdom. Their genomes carry single nucleotide polymorphisms and deletions that are characteristic of the B.1.1.7 variant of concern, but lack the full complement of lineage-defining mutations. Instead, the remainder of their genomes share contiguous genetic variation with non-B.1.1.7 viruses circulating in the same geographic area at the same time as the recombinants. In four instances there was evidence for onward transmission of a recombinant-origin virus, including one transmission cluster of 45 sequenced cases over the course of two months. The inferred genomic locations of recombination breakpoints suggest that every community-transmitted recombinant virus inherited its spike region from a B.1.1.7 parental virus, consistent with a transmission advantage for B.1.1.7’s set of mutations.

## Introduction

Recombination, the transfer of genetic information between molecules derived from different organisms, is a fundamental process in evolution because it can generate novel genetic variation upon which selection can act (reviewed in Felsenstein 1974). Genetic analysis indicates that recombination occurs frequently in betacoronaviruses (Lai et al. 1985; Keck et al. 1988; Lai and Cavanagh 1997), including natural populations of MERS-CoV (Corman et al. 2014; Dudas and Rambaut 2016; Kim et al. 2016) and SARS and SARS-like coronaviruses (Hon et al. 2008; Boni et al. 2020). It has been proposed recently that the global SARS-CoV-2 genome sequence data contains signals of recombination across the pandemic (VanInsberghe et al. 2021). Recombination has the potential to be important in the context of pathogen evolution because it can “rescue” genomes with otherwise deleterious mutations or provide the opportunity to create novel phenotypes by bringing genetic variation from different backgrounds onto a single genome. A concerning scenario from an epidemiological perspective is the potential for recombination to combine, in the same genome, mutations that may confer immune-escape properties with those that may enhance transmissibility. Enhanced transmissibility (Volz et al. 2021) and immune-escape (Planas et al. 2021) phenotypes have already been observed in SARS-CoV-2. Consequently, the characterization of recombination in SARS-CoV-2 is important for surveillance purposes.

The molecular mechanism of homologous recombination in unsegmented positive-sense RNA viruses such as SARS-CoV-2 is generally by copy-choice replication, a model first suggested in poliovirus (Cooper et al. 1974). In this process a hybrid or mosaic RNA is formed when the RNA-polymerase complex switches from one RNA template molecule to another during replication (Worobey and Holmes 1999). In order for homologous recombination to occur, and be subsequently detected, there must be co-infection of the same cell within an individual by genetically-distinct viruses (termed the ‘parental’ lineages of the recombinant virus). Co-infection of an individual requires there to be co-circulation of multiple viral lineages within a population and, given the short duration of most SARS-CoV-2 infections, is most likely to be observed when virus prevalence is high.

Conditions conducive to SARS-CoV-2 recombination existed in the United Kingdom (UK) during the latter part of 2020 and early in 2021. From mid-October 2020 to January 2021, SARS-CoV-2 prevalence was estimated to be between 1 and 2% in England (Steel and Hill 2021). During this time, the B.1.1.7 variant of concern (VOC) emerged, rapidly increased in frequency, and spread across the UK, replacing lineages that were already at high prevalence (Volz et al. 2021). The most common of the latter was the B.1.177 lineage and its descendants (Hodcroft et al. 2020) (Figure 1). B.1.1.7 is characterised by an unusually large number of genetic changes (22 mutations from its immediate ancestor; Rambaut et al. 2020a). The ability to detect virus recombination using comparative sequence analysis depends on the genetic distinctiveness of the parental viruses, so the co-circulation of B.1.1.7 and non-B.1.1.7 viruses is expected to increase the power to detect recombinants between these lineages. The UK’s high rate of genomic surveillance and unified collection of genomic, epidemiological, and geographic data also provide multiple lines of evidence for evaluating the identification of recombinant viruses.

**Figure 1.**
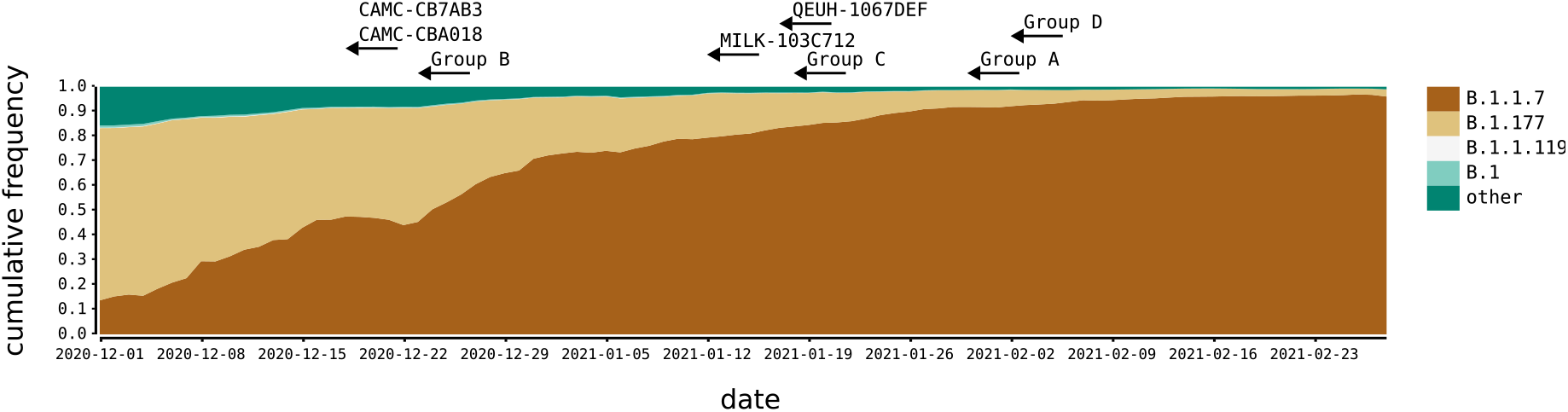
SARS-CoV-2 lineages in the UK, Winter 2020-2021. The distribution of the most frequent SARS-CoV-2 lineages in the United Kingdom from December 2020 to February 2021. Here, B.1.177 refers to B.1.177 including all of its descendant lineages (e.g. B.1.177.9). For each recombinant, or recombinant group, the date of the earliest sampled genome is indicated by the left tip of the arrows. The recombination event that generated each must have occurred before this date.

To identify putative SARS-CoV-2 recombinant viruses we carried out an analysis of all complete UK SARS-CoV-2 genomes that had been assigned to lineage B.1.1.7 and which showed evidence of being the product of combining different genetic lineages, indicative of recombination. Specifically, we scanned the UK dataset for genomes that were alternately composed of long contiguous tracts of B.1.1.7 and non-B.1.1.7 genetic variation. The genetic composition and epidemiological context of each candidate mosaic genome was carefully explored to determine whether it was recombinant in origin. We subsequently determined whether the recombinants showed evidence of onward transmission within the UK population. One recombinant lineage continued to circulate for at least nine weeks and, as of 5th May 2021, was associated with 45 linked infections.

## Results

### Identification of putative recombinants

We identified a total of 16 recombinant sequences from the whole UK dataset of 279,000 sequences up to the 7th March 2021, using our bioinformatic and evolutionary analysis pipeline (see Methods). Twelve genome sequences that clustered into four groups (labeled A - D) and four additional singletons showed evidence of being mosaic in structure (Table 1; Supplementary Table S1). For each group A-D, each of the constituent genomes was sampled from the same geographic locality within the UK (Table 1). For Group A, which spanned two geographical regions, all the samples originated from close to the border between Wales and the North-West of England (<20km apart). The sample dates for the putative recombinants ranged from 2020/12/18 - 2021/02/02 (Figure 1). If Groups A-D represent community transmission of a recombinant lineage with a single origin, recombination must have occurred on or before the date of the earliest sample in each group. This range coincides with a period of increasing relative prevalence of B.1.1.7 in the UK alongside the presence of other circulating lineages in the community, the most common of which were B.1.177 and its descendants (Figure 1).

**Table 1.** Recombinants and their putative second parental lineages according to genetic similarity (the first parental lineage is always B.1.1.7). For the recombinant groups the number of genomes, the NUTS1 location of residence and the range of sampling dates are given. Breakpoint coordinates are the range of possible SARS-CoV-2 genome positions bounded by mutations that are unambiguously inherited from one parent or the other, including both single nucleotide polymorphisms and deletions. The date and location for the 2nd parent is for the genetically most similar UK genome(s) within the genome region belonging to that lineage. †For Group A, the results for LIVE-DFCFFE (in parentheses) were different to those of the rest of the group.

|  | No. | Location | Sample dates | 2<sup>nd</sup> parent lineage | 2<sup>nd</sup> parent date | 2<sup>nd</sup> parent location | Breakpoint coordinates |
| --- | --- | --- | --- | --- | --- | --- | --- |
| Group A | 4 | Wales / North West England | 2021/01/30 – 2021/02/21 | B.1.177 | 2021/01/27 | Wales | 21255 – 21765 (20410 - 21765)<sup>†</sup> |
| Group B | 2 | South East England | 2020/12/23 – 2020/12/24 | B.1.36.28 | 2020/12/10 | Greater London | 6528 – 6954 |
| Group C | 3 | East Midlands (England) | 2021/01/18 – 2021/01/30 | B.1.221.1 / B.1.221.2 | 2021/01/02 | East Midlands / West Midlands / Greater London | 24914 – 28651 |
| Group D | 3 | South East England | 2021/02/02 – 2021/02/07 | B.1.36.17 | 2020/12/10 | East of England | 21575 – 23063 |
| CAMC-CBA018 | 1 | Greater London | 2020/12/18 | B.1.177 | 2021/01/23 | UK | 11396 – 21991 |
| CAMC-CB7AB3 | 1 | Greater London | 2020/12/18 | B.1.177 | 2020/12/12 | Greater London | 3267 – 6286 & 12534 – 21765 |
| MILK-103C712 | 1 | Greater London | 2021/01/12 | B.1.177.16 | 2020/12/14 | Greater London / East of England | 26801 – 27972 |
| QEUH-1067DEF | 1 | Scotland | 2021/01/17 | B.1.177.9 | 2021/01/13 | Greater London | 6954 – 10870 |

To rule out the possibility that any of the sixteen recombinants could have resulted from artefacts as a result of assembling sequence reads from a co-infected sample (generated through either natural co-infection or laboratory contamination), we examined the read coverage and minor allele frequencies and assessed the likelihood of a mixed sample. Several lines of evidence suggested the recombinant sequences were not the products of sequencing a mixture of genomes: Firstly, the sequencing protocol used in the UK (Tyson et al. 2020) generates 98 short (∼350bp) amplicons, such that long tracts that match just one lineage would be unlikely. Secondly, the read data do not support a mixture for any of the putative recombinant genomes. All the recombinants were sequenced to high coverage (lowest mean read depth per site per genome: 686; highest mean read depth: 2903). The mean minor allele frequency (MAF) for the putative recombinants was 0.008, which is 6 standard deviations below the mean of the MAF (0.34) for a set of 20 sequences that we suspected to be mixtures (Supplementary Figure S1). Finally, for all groups A-D, multiple genomes with the same mosaic structure were sequenced independently from different samples, and by different sequencing centres in the case of Group A, implying that the original assembly was correct and, additionally, that transmission of the recombinant had occurred. All of the read data are available on the European Nucleotide Archive. Accession numbers are given in Supplementary Table S2.

### Epidemiological information supports the identification of putative parental lineages

The nucleotide variation for the putative recombinants and their closest neighbours by genetic similarity (for each of the regions of their genomes either side of the recombination breakpoint) is shown in Figure 2 and Supplementary Figure S2. For each of Groups A-D, the closest neighbours by genetic similarity for each of the two non-recombining genome regions were the same sequences for every putative recombinant within a group. For most of the recombinants, there were several equidistant putative parental sequences for each region of the genome; whenever this was true, they all belonged to the same lineage, except for Group C, whose putative parental lineages for the non-B.1.1.7-like region of the genome were a mixture of two closely related lineages (B.1.211.1 and B.1.211.2). The putative parental sequence for the non-B.1.1.7 region of the genome varied by group (Table 1). Importantly, in each case, the sequence and epidemiological data demonstrate that the non-B.1.1.7 parental sequence was circulating in the same geographic area as the recombinant in the time immediately before the sampling date of the recombinant. For Group A and the four singletons, the second parental sequence was assigned lineage B.1.177 or one of its descendants. B.1.177, which likely arose in Spain in the summer of 2020 and was exported to multiple European countries (Hodcroft et al. 2020), rose to high relative frequency in the UK through Autumn 2020, and was widespread by December (Figure 1). Lineage B.1.177.16, the second parental sequence of MILK-103C712, was sampled 25 times in Greater London in the four weeks preceding MILK-103C712’s sample date. Lineage B.1.177.9 was sampled on one other occasion in Scotland in four weeks preceding QEUH-1067DEF’s sample date. Lineage B.1.36.28 was not sampled in the South-East of England in the four weeks preceding Group B’s sample date, but was sampled eight times in Greater London in that period. Lineages B.1.221.1 and B.1.221.2 were sampled seven and zero times, respectively, in the East Midlands in the four weeks preceding Group C’s sample date, and B.1.36.17 was sampled five times in the South East in the four weeks prior to Group D’s sample date. The distributions of the most prevalent lineages over time in each region of the UK relevant to the eight sets of recombinants (Groups A-D and the four singletons) are shown in Supplementary Figure S3.

**Figure 2.**
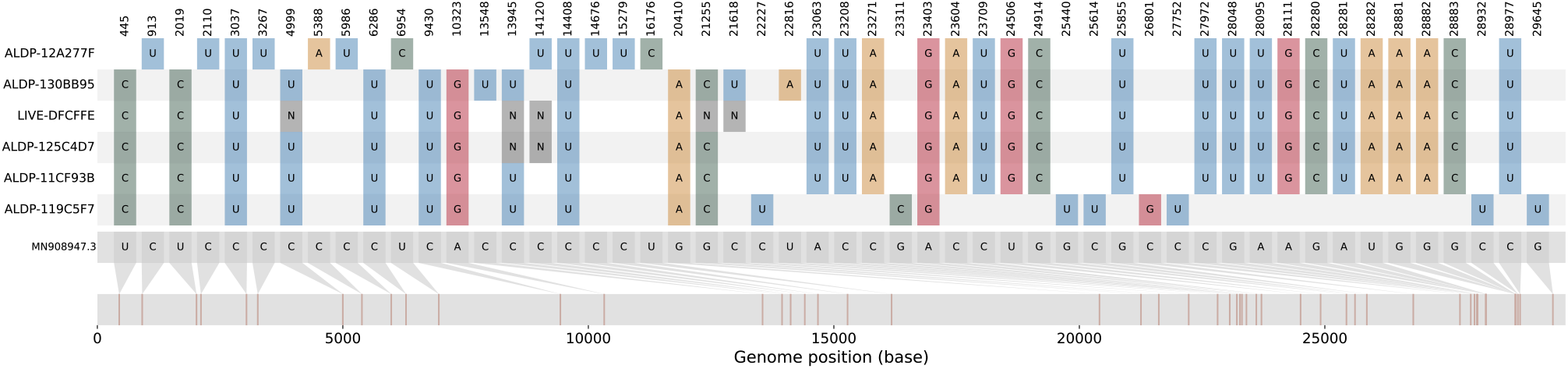
The nucleotide variation present in Group A. The nucleotide variation with respect to the reference sequence (MN908947.3; grey genome far bottom) for the four members of Group A (ALDP-11CF93B, ALDP-125C4D7, LIVE-DFCFFE, ALDP-130BB95; middle four coloured genomes) and their closest neighbours by genetic similarity among all UK sequences from the same time period for the B.1.1.7-like region of their genomes (ALPD-12A277F; top coloured genome) and the B.1.177-like region of their genomes (ALDP-119C5F7; bottom coloured genome).

### Putative recombinants exhibit significant mosaicism

We rejected the null hypothesis of non-reticulate evolution for 14 out of the 16 putative recombinant sequences by testing these sequences for mosaicism (3SEQ (Lam, Ratmann, and Boni 2018) with Dunn-Sidak correction for multiple comparisons) against a background set of 2000 sequences randomly drawn from the course of the UK epidemic (Table 2). The lineages identified as the putative parentals assigned by 3SEQ agreed with the lineages for putative parentals assigned by genetic similarity (Tables 1 and 2) even though of the 16 closest neighbours by genetic similarity described above, none were present in the background sequence set of candidate parentals used in the 3SEQ analysis. The breakpoints reported by 3SEQ also agreed with breakpoints inferred from the distribution of Single Nucleotide Polymorphisms (SNPs) and deletions in the putative recombinants and their neighbours by genetic similarity (Tables 1 and 2). The two sequences that belong to Group B did not show a statistically significant mosaic signal of non-reticulate evolution, but 3SEQ’s Δ_m,n,2_ statistic for these two candidate recombinants showed the greatest support for mosaicism possible among the ancestry-informative polymorphic sites with their closest neighbours by genetic similarity as parentals: n = 6, m = 42, k = 42. The associated uncorrected p-value of 5.7e-7 does not survive a multiple comparisons correction due to the number of putative parental lineages and descendants that were tested (Table 2)

**Table 2.** Recombinants, their putative second parental lineages, and inferred breakpoints according to 3SEQ (the first parental lineage is always B.1.1.7). P-values for the Δ_m,n,2_ statistic are reported uncorrected, and after Dunn-Sidak correction for multiple comparisons. * Statistics are from a separate run with the closest neighbours by genetic distance included in the parental dataset. † For Group A, the results for LIVE-DFCFFE (in parentheses) were different to those of the rest of the group.

|  | 2<sup>nd</sup> parent lineage | Uncorrected p-value | Corrected p-value | Breakpoint coordinates |
| --- | --- | --- | --- | --- |
| Group A | B.1.177 | 2.15E-12 (9.83E-13)<sup>†</sup> | 0.00003 (0.00006)<sup>†</sup> | 21255-21613<br>(18998-20294)<sup>†</sup> |
| Group B* |  | 5.70E-07 | 1 | 6528 – 6954 |
| Group C | B.1.221.1 | 4.267E-10 | 0.02686 | 25996-27441 |
| Group D | B.1.36.39 | 3E-12 | 0.002 | 20703-23063 |
| CAMC-CBA018 | B.1.177 | 2E-12 | 0.00014 | 20389-21255 |
| CAMC-CB7AB3 | B.1.177 | 1E-11 | 0.00065 | 3267-4474 & 20389-21254 |
| MILK-103C712 | B.1.177.17 | 1.06E-10 | 0.00673 | 408-444 & 26801-27876 |
| QEUH-1067DEF | B.1.177.9 | 4.12E-10 | 0.026 | 10523-10870 |

The Dunn-Sidak correction used in 3SEQ is very conservative as it assumes that all 64.0 million comparisons we performed are independent statistical tests, when in fact these tests are highly non-independent since many candidate parental sequences are a small number of nucleotide differences apart from each other. When corrected p-values are borderline, the recommended approach to infer non-reticulate evolution is to build separate phylogenetic trees for the non-recombining regions of the genome to confirm that the recombinant in question has different phylogenetic placements in different genomic regions (Boni et al. 2010). With the exception of the inner region for CAMC-CB7AB3, whose placement within B.1.177 was not well supported, each recombinant’s two phylogenetic placements were with the lineages that we identified as parental by genetic similarity and by using 3SEQ, with high bootstrap support (Supplementary Table S3). The placement of the two parental genome regions for each recombinant in the context of the whole epidemic in the UK is shown in Figure 3.

**Figure 3.**
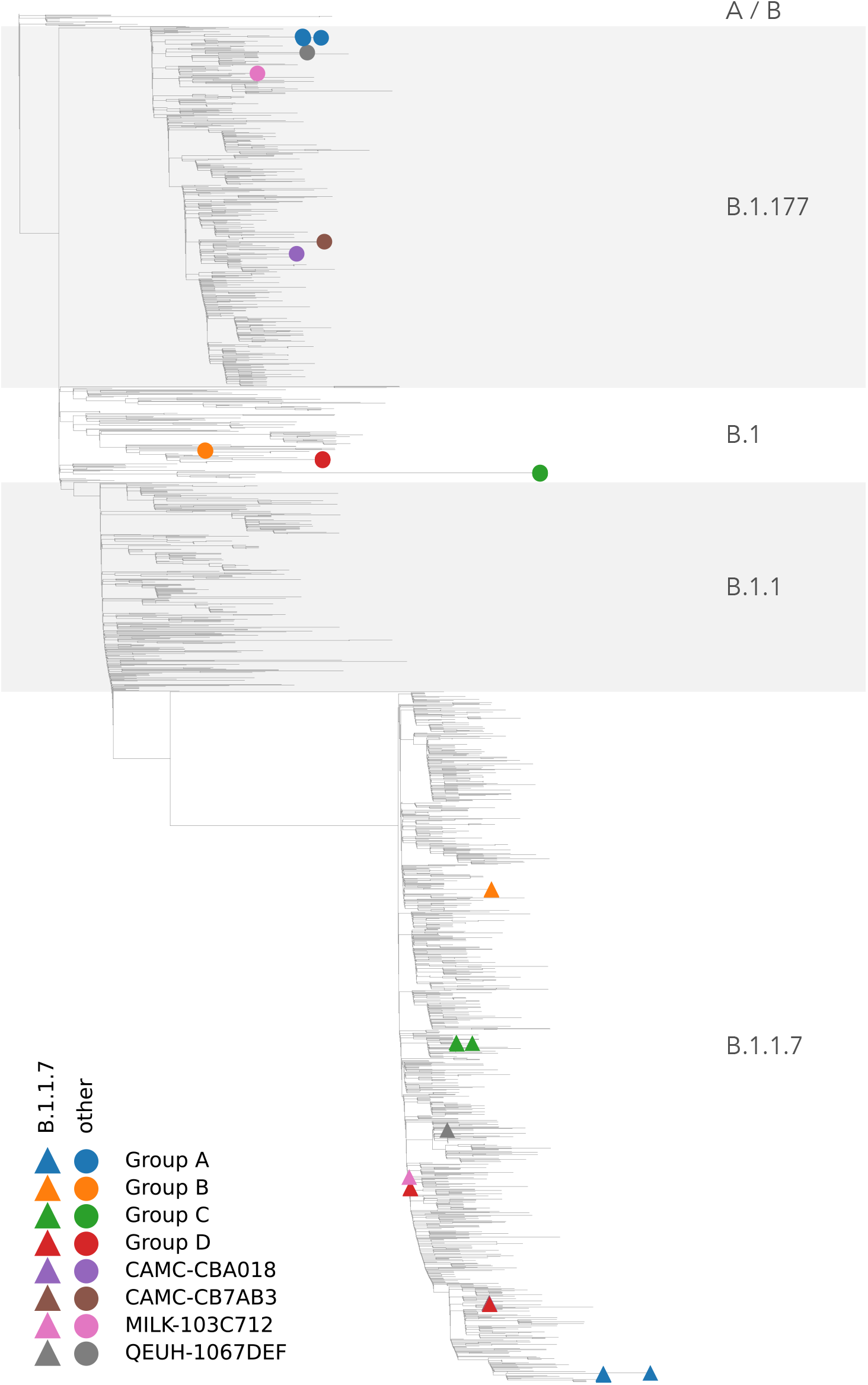
Phylogenetic placement of putative recombinant genome regions. Phylogenetic reconstruction of 2000 samples chosen to be representative of the course of the epidemic in the UK, as well as the 16 recombinant genomes, with their B.1.1.7-like part (coloured triangles) and non-B.1.1.7-like part (coloured circles) alternately unmasked.

The mosaic structures of the genomes of the putative recombinants are shown in Figure 4. In six out of eight instances (and all four of the groups of >1 sequence, which may represent community transmission), the recombinants contain a spike gene from the B.1.1.7 lineage. In four instances there is a proposed recombination breakpoint at or near the 5’ end of the spike gene.

**Figure 4.**
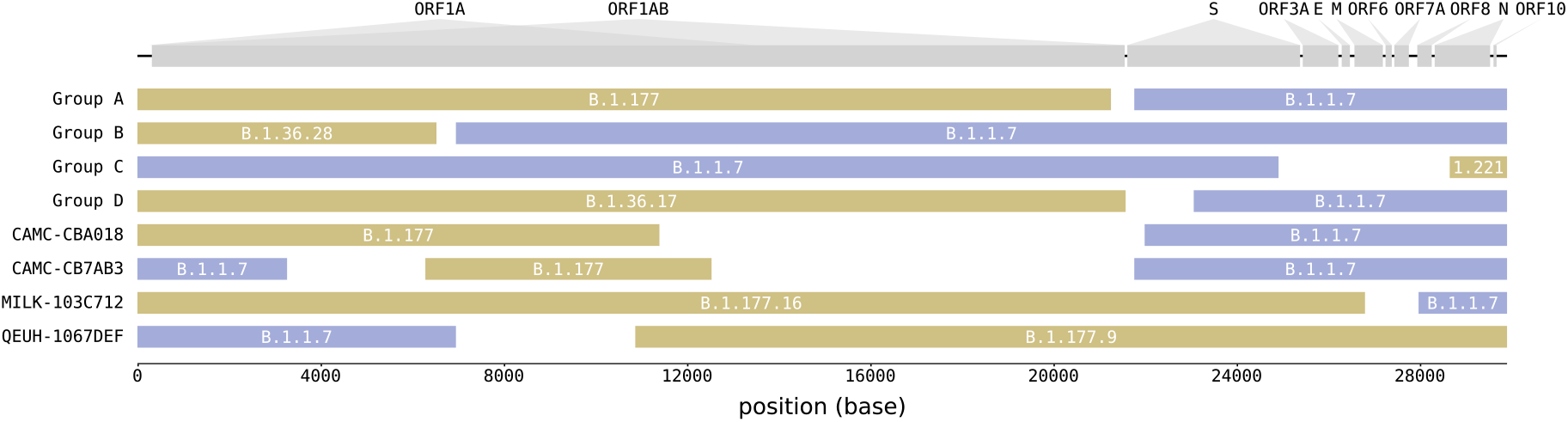
Mosaicism of putative recombinants. Recombinant groups A-D contain multiple sequences exhibiting the same mosaic genome structures (see Table 1 for details). Tracts matching lineage B.1.1.7 are shown in blue, whilst virus genome regions matching other lineages are shown in yellow. Gaps represent ambiguity in the exact position of the recombinant breakpoints; there are no lineage-defining mutations within these regions. The breakpoint coordinates are taken from Table 1.

### Further evidence for the community transmission of Group A

A follow-up investigation of the eight sets of putative recombinants (Groups A-D and the four singletons) on 5th May 2021 found 41 sequences that were descended from Group A (Figure 5). No descendants from any other recombinant event were detected to have continued to circulate. The 41 sequences share the same set of SNPs with three members of Group A (ALDP-11CF93B, ALDP-125C4D7 and LIVE-DFCFFE), with additional nucleotide variation at positions 8090, 16260 and 25521 (Figure 5A; Supplementary Figure 4). They were sampled in the North West of England, between 2021/03/01 and 2021/04/04 (Figure 5B; C). The temporal distribution of samples descended from the recombination event that led to Group A suggests that the recombinant lineage persisted at low frequency for a period of time before expanding and then contracting again (Figure 5C). A second follow-up investigation on the 1st June found no further recombinants descended from Group A, which suggests that this transmission cluster is extinct. The dynamics of this cluster of infections reflect the wider trend of SARS-CoV-2 prevalence in England over the same time period. Prevalence decreased from a maximum of 2.08% in January 2021 to 0.21% for the week beginning 2021/04/04 and to <0.1% as of the beginning of May, 2021 (Steel and Hill 2021). Group A and its descendants met the criteria for designation as a recombinant Pango lineage (Pybus 2021), and have been named lineage XA (https://cov-lineages.org/lineages/lineage_XA.html).

**Figure 5.**
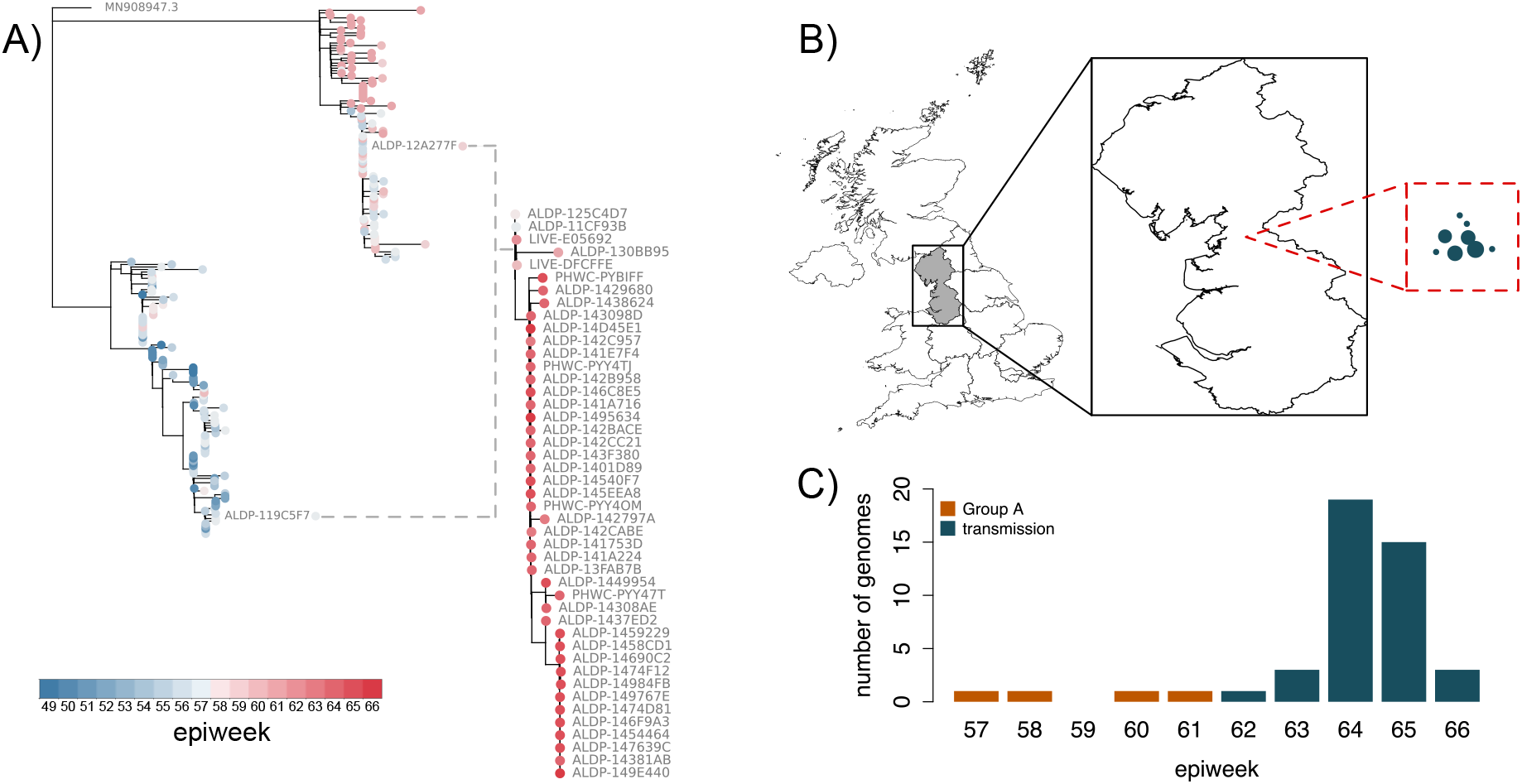
The community transmission of Group A. A) The phylogenetic relationships within Group A and their descendants (right-hand tree) and their closest neighbours by genetic similarity for the B.1.1.7-inherited region of their genome (top clade; left-hand tree) and the B.1.177-inherited region of their genome (bottom clade; left-hand tree). The branch lengths are not plotted to the same scale for the two trees. The closest parental sequences by genetic similarity for the two regions of the genomes (ALDP-12A277F and ALDP-119C5F7) are highlighted in the left-hand tree. The sample date in cumulative epidemiological weeks (epiweeks) since the first epiweek of 2020 for each sequence is represented by coloured circles at the tips of each tree. The dashed lines represent the formation of a new recombinant clade between the members of Group A and their parental lineages. B) The geographic context of the transmitted recombinant sequences. The exploded region of the map is the North West region of England. All of the 41 recombinants descended from Group A were sampled in this region. The relative distribution of their locations, in the same scale as the exploded region, are represented by the circles in the red dashed square. The size of the points represents the number of genomes sequenced in each location. The absolute locations of the recombinants within North West England are not represented by this panel. C) The distribution of the sampling dates for the 45 recombinants, aggregated by epiweek. Orange bars: the four original members of Group A; Green bars: the 41 descendants from Group A.

## Discussion

Here we report the first unambiguous detection and characterisation of the arisal and subsequent community-transmission of recombinant SARS-CoV-2 viruses. Comparison of intra-genomic variation, supported by geographic and epidemiological data, demonstrates the occurrence of multiple independent recombination events involving UK virus lineages in late 2020. Recombinant genomes that share genetic identity were sampled from the same geographic location and time period, indicating they represent successful onward transmission after the occurrence of a single ancestral recombination event. In one instance this resulted in a significant transmission cluster comprising 45 observed cases, which has been given the Pango lineage name XA. While no obvious biological advantage can be attributed to this cluster (or to any of the observed recombinants) beyond the acquisition of B.1.1.7’s set of spike mutations, these recombinants are sentinel events for continued monitoring for new variants. With the increasing co-circulation of variants of concern in the same geographic areas careful monitoring is warranted.

Large-scale bioinformatic approaches have identified statistical signals of recombination among SARS-CoV-2 sequences, using clade assignment and its changes along the genome as the primary characteristic under investigation (Varabyou et al. 2020; VanInsberghe et al. 2021). Due to the limited genetic diversity at the time these analyses were carried out, there was no strong statistical support for recombination (as opposed to non-reticulate diversification) for any particular candidate recombinant. When the number of mutations in a virus sequence is low (e.g. Figure 3 in VanInsberghe et al. (2021); Figure 2 in Varabyou et al. (2020)) there is generally little statistical support to reject the possibility that the sequence patterns could have been generated by mutation alone. In contrast, there is sufficient diversity in the UK virus dataset between the lineage B.1.1.7 and other co-circulating lineages to detect putative recombinants and demonstrate statistical significance for their breakpoint patterns. Candidate recombinants and their parental lineages are a median of 22.5 nucleotide mutations apart; and candidate pairs of parentals are a median of 46 nucleotide differences apart from each other (Figure 2, Supplementary Figure S2). The p-values we present are exact non-parametric probabilities that the observed patterns of nucleotide ancestry in the candidate recombinant viruses were generated by mutation alone given the parental genotypes, and are corrected for multiple testing.

The breakpoint locations inferred from the recombinants’ parental lineages by two different methods and two different parental datasets are in agreement (cf. Tables 1; 2). Two interesting observations arise from their distribution. Firstly, in six cases, and in all the cases where we detected transmission, the spike gene was inherited from the B.1.1.7 parental sequence (Figure 4). This is consistent with the observed transmission advantage of B.1.1.7 (Volz et al. 2021), which is likely attributable to the mutations it carries in the spike region (Rambaut et al. 2020a). Secondly, in four instances a breakpoint is located near the 5’ end of the spike gene. A defining feature of the Order *Nidovirales*, to which coronaviruses belong, is the production during RNA synthesis of a set of nested positive- and negative-stranded subgenomic RNAs (sgRNAs) that contain a leader sequence derived from the 5’ end of the complete genome and a progressively reduced complement of the structural (S, E, M, N) and accessory genes, which form the body of the sgRNA molecule (Masters 2006; Kim et al. 2020). The discontinuous nature of these sgRNAs is understood to be the product of template switching by viral polymerase during normal transcription, where the polymerase pauses at a transcription-regulatory sequence (TRS) after transcribing the last open reading frame (ORF) of the sgRNA, and switches to a similar TRS upstream of the leader sequence (Sawicki and Sawicki 1995), omitting a looped-out region of the template RNA, which contains at least orf1ab in the case of SARS-CoV-2 (Finkel et al. 2021). This provides an environment that is highly conducive to homologous recombination: a polymerase that engages in template switching during its normal transcriptional activity, as well as the availability of alternative template RNA molecules, in the form of sgRNAs, which incorporate sequence motifs that mediate template switching, and which might be derived from different genomes in the case of coinfection. As TRSes, which occur between the ORFs (Kim et al. 2020), are important in mediating template switching during homologous recombination in coronaviruses, this can account for the shared pattern of recombination-prone regions observed here (Figure 4). However, to be detected recombinant genomes must lead to viable viruses, so the distribution of breakpoints observed from genomic surveillance may not represent the distribution of breakpoints that occur in situ (Banner and Lai 1991).

Given the overall prevalence of SARS-CoV-2 in the UK in December 2020 of 1% (Steel and Hill 2021) and assuming that 40% of infections were of lineage B.1.1.7 and 40% of B.1.177 (Figure 1), a simplistic expectation for the number of co-infections involving these two lineages is the product of their prevalences, which is 16 co-infections per million people. The figure is 70 co-infections per million people for a prevalence of 2%, 70% infection with B.1.1.7 and 25% infection with B.1.177, which applies to the first half of January 2021. In this calculation we assume that infections are independent and that once infected the chance of an additional infection occurring is unchanged. This illustrates that on a national scale, co-infection should be quite common during periods of high prevalence. From a public health perspective, this reminds us that halving prevalence reduces the chance of coinfection by a factor of four, because the probability of coinfection increases with the square of the prevalence.

As recombination permits the combination of advantageous mutations from distinct variants, and recombination is only possible with co-infection, minimising the prevalence of SARS-CoV-2 will minimize the chance of forming recombinant lineages with genetic combinations that could potentially increase virus fitness. At the time of writing, the trajectory of the SARS-CoV-2 pandemic continues to remind us that there are many populations worldwide still highly susceptible to large epidemic waves. High prevalence epidemic waves comprising diverse viral lineages risk high rates of meaningful recombination, and as SARS-CoV-2 genetic diversity is much greater in 2021 than in 2020, it will be important to examine each epidemic for the presence of novel recombinant lineages, especially epidemics occurring in regions with circulation of different variants of biological significance.

## Supporting information

COG-UK Authors

## Data Availability

## Resource availability

## Lead Contact

Further information and requests for resources and reagents should be directed to and will be fulfilled by the Lead Contact

## Materials Availability

This study did not generate new unique reagents.

## Data and code availability

All the code used to perform the analyses here is available at: https://github.com/COG-UK/UK-recombination-analysis

## Acknowledgements

COG-UK is supported by funding from the Medical Research Council (MRC) part of UK Research & Innovation (UKRI), the National Institute of Health Research (NIHR) and Genome Research Limited, operating as the Wellcome Sanger Institute. OGP was supported by the Oxford Martin School. JTM, RMC, NJL and AR acknowledge the support of the Wellcome Trust (Collaborators Award 206298/Z/17/Z – ARTIC network). DLR acknowledges the support of the MRC (MC_UU_12014/12) and the Wellcome Trust (220977/Z/20/Z). ES and AR are supported by the European Research Council (grant agreement no. 725422 – ReservoirDOCS). TRC and NJL acknowledge the support of the MRC, which provided the funding for the MRC CLIMB infrastructure used to analyse, store and share the UK sequencing dataset (MR/L015080/1 and MR/T030062/1). The samples sequenced in Wales were sequenced partly using funding provided by Welsh Government.

## Author contributions

Conceptualization: AR, MFB, OGP,

BJ Data curation: MJB, NP

Formal analysis: BJ

Funding acquisition: AR, TRC, NJL, OGP

Resources: TRC, SN, RP, AC, AD, SH, AL, NP, FT, HW, MW, CW

Software: MFB, AO’T, ES, BJ, MJB

Visualization: BJ, AO’T

Writing - original draft: BJ, MFB, OGP, AR, DLR

Writing - review and editing: MFB, DLR, OGP, TRC, AR

## Resource availability

### Materials Availability

This study did not generate new unique reagents.

## METHOD DETAILS

### Identification of putative recombinants

A national SARS-CoV-2 sequencing effort in the UK, the COG-UK consortium (COVID-19 Genomics UK (COG-UK) 2020), has undertaken systematic genomic surveillance of SARS-CoV-2 in the country and generated over 440,000 genomes to date. As part of the COG-UK daily analytical pipeline (https://github.com/COG-UK/grapevine_nextflow), the consensus genome sequences of the complete set of UK samples were aligned to the SARS-CoV-2 reference sequence (Genbank accession: MN908947.3) using Minimap2 (Li 2018). The aligned sequences were converted from sam to fasta format, and each assigned a Pango lineage (Rambaut et al. 2020b) using Pangolin (https://github.com/cov-lineages/pangolin). Pango lineages are designed to capture ongoing epidemiological trends at a resolution suitable for genomic epidemiology and outbreak investigation (Rambaut et al. 2020b). From the sequence alignment, we extracted all sequences that had been assigned to lineage B.1.1.7, up to the 7th March 2021. We genotyped these sequences at the set of 22 sites that discriminate B.1.1.7 from its parental lineage (B.1.1) using a custom script in Python (https://github.com/cov-ert/type_variants), then discarded sequences with missing data at any of the 22 sites. We visualised the resulting table of genotype calls in order to identify sequences that showed evidence of a potential mosaic genome structure (i.e. runs of contiguous sites that were not compatible with the B.1.1.7 lineage designation).

### Identification of candidate parental sequences

To identify candidate parental genome sequences in a computationally-tractable manner we created a set of all UK SARS-CoV-2 sequences that (i) contained no N nucleotide ambiguity codes after masking the 3’ and 5’ UTRs, (ii) spanned the dates 01/12/2020 to 28/02/2021, which represents two weeks before the date of the earliest putative recombinant, to one week after the date of the latest, and (iii) excluded the putative recombinant genomes identified above. This set consisted of 98859 sequences in total. For each putative recombinant, we split its genome sequence into B.1.1.7-like regions and non-B.1.1.7 regions at the junction of genetic regions according to the mosaic structure detected by the custom Python script described above (https://github.com/cov-ert/type_variants; Supplementary Table S1). Then for each component region of each mosaic genome, we first masked the remainder of the genome with Ns (in both the focal mosaic sequence and all background sequences) then found the most-genetically similar non-focal sequences by computing pairwise genetic distances (number of nucleotide differences per site) using gofasta (https://github.com/cov-ert/gofasta). Subsequently, an alignment was compiled for each putative recombinant, which contained the putative recombinant as well as the most-genetically similar background sequences (as identified above) for each component region of that mosaic genome. The single nucleotide differences between the putative recombinant and the closely related reference sequences were visualised using snipit (https://github.com/aineniamh/snipit). The genomic coordinates of the boundaries between each mosaic genome region were then refined by taking into account observed lineage-defining nucleotide and deletion variation. Specifically, we set the boundary coordinates to the ends of sequential tracts of mutations specific to the putative parental sequences. This is a conservative approach to assigning parental lineages and consequently no parental lineage is assigned to those genome regions that do not contain unambiguous lineage-defining mutations or deletions. Lastly, using these refined region boundaries, we reiterated the genetic distance calculation above to identify a final set of most-genetically similar sequences for each putative recombinant.

When reporting geographic locations for UK virus genome sequences we use level 1 of the Nomenclature of Territorial Units for Statistics (NUTS) geocode standard (https://ec.europa.eu/eurostat/web/nuts/history).

### Defining a representative sample from the UK epidemic

To generate a limited set of genomes that are suitable for computationally-expensive analysis yet are also representative of the genetic diversity of the SARS-CoV-2 epidemic in the UK, we randomly sampled 2000 sequences from 21st March 2020, when sequence data first became available, to 1st March 2021, weighting the probability of choosing a sequence accounting for the sequencing coverage and covid19 prevalence in individual geographic regions of the UK over time, using the same method as in (Volz et al. 2021), which is available at (https://github.com/robj411/sequencing_coverage). We use this dataset to investigate the phylogenetic placement of the alternate regions of recombinant genomes, and as a dataset of putative parental sequences to statistically test for recombination using 3SEQ.

### Investigation of read data

Almost all sequencing sites in the COG-UK consortium use the ARTIC PCR protocol to produce tiled PCR amplicons, which are then sequenced (Tyson et al. 2020). The generated sequence reads are then processed using sequence mapping, rather than sequence assembly, to produce a consensus genome for each sample. This approach, which was designed to support epidemiological investigations, creates a single consensus sequence for each sample. Beyond representing sites with high minor allele frequencies using the appropriate IUPAC nucleotide alphabet ambiguity code, this consensus does not reflect the natural genetic variation of SARS-CoV-2 genomes observed within an infected individual (Lythgoe et al. 2021). Mapping is particularly suited to tiled amplicons generated from samples that contain limited genomic diversity. Further, mapping is typically less prone to introducing errors/artefacts than sequence assembly and enables effective primer sequence removal and identification of non-reference mutations. Genomic sites that exhibit intra-sample nucleotide variation could be consistent with a range of processes, including co-infection, within-patient diversity, contamination, or PCR error. The identification of such sites forms part of the consensus-generating pipeline, and we exploit that information here in order to rule out the possibility that our mosaic consensus sequence represents a mixture of virus genomes, rather than representing true recombinant genomes.

For each putative recombinant sequence, we analysed the original read data from virus genome sequencing in order to rule out the possibility that the generated consensus sequence represents a mixture of virus genomes (due to laboratory contamination or coinfection, for example), rather than representing a true recombinant genome. To do this we calculated minor allele frequencies (MAFs) from the read data and compared their distribution between the 16 recombinant genomes and 20 samples that we suspected of being the product of sequencing a mixture of genomes, potentially because of coinfection or laboratory contamination. To define sequences that we suspected of being mixtures, we scanned the dataset for consensus sequences that possessed an IUPAC ambiguity code at the 27 genomic positions that differ from the SARS-CoV-2 reference genome (Genbank accession: MN908947.3) by a nucleotide change in B.1.1.7 (the 27 positions include those with nucleotide changes that were inherited from the ancestor of B.1.1.7). We define the MAF at a single site as the number of sequencing reads not containing the most frequently observed single nucleotide allele that mapped to that site, divided by the total number of sequencing reads that include any nucleotide allele that mapped to that site. For each virus genome, we defined a set of genomic positions from which to calculate MAF as follows. For each recombinant, we considered every site that differed from MN908947.3 by a nucleotide in its own consensus genome, or in the consensus genome of either of its parentals by genetic similarity. For the sequences that we suspected of being mixtures we considered the 27 genomic positions where sequences belonging to B.1.1.7 differ from MN908947.3 by a nucleotide change. We used samtools (Li et al. 2009), with default filters for mapping and base quality, to extract allele calls from the read data using its mpileup subroutine, and to calculate mean read depth per genome using its depth subroutine.

## QUANTIFICATION AND STATISTICAL ANALYSIS

### Test for mosaic genome structure

We used 3SEQ (Lam, Ratmann, and Boni 2018) as a statistical test for recombination in the UK SARS-CoV-2 data. 3SEQ interrogates triplets of sequences for a signal of mosaicism in one “child” sequence, given the genotypes of the other two “parental” sequences, using an exact non-parametric test for clustering in a sequence of binary outcomes (Boni, Posada, and Feldman 2007). The test statistic Δ_m,n,2_ used in 3SEQ simply tests if a putative recombinant’s ancestry in parental A clusters in the middle of the genome, while ancestry in parental B clusters in the outer regions of the genome. We manually adjusted two-breakpoint recombinants to be single-breakpoint recombinants if one of the breakpoints according to 3SEQ abutted the beginning or end of the genome. We tested all potential pairs of sequences from the representative parental dataset from the course of the UK pandemic (n = 2000) against each putative recombinant in the child dataset (n = 16), and report p-values that are uncorrected and that are Dunn-Sidak corrected for multiple comparisons (n = 64.0 million). We performed a single additional run of 3SEQ with two putative recombinant sequences that were not found to be significantly the mosaic product of any of the sequences in the representative background as children, and their closest neighbours by genetic similarity as parentals. P-values for this test were reported without correction and after correction for multiple testing assuming that this test was in addition to the 64 million comparisons that we had already performed.

### Test for the phylogenetic incongruence of putative recombinant genome tracts

For each of the eight sets of recombinants (Groups A-D and the four singletons) we carried out the following procedure to test for incongruence between the phylogenetic placements of the two regions of their genomes. We independently added each set’s genome(s) to the representative background of 2000 sequences, along with the reference sequence, to create eight alignments in total. We masked the resulting alignments according to the breakpoints defined by the closest neighbours by genetic similarity, so that for each set, we produced two sub-alignments: one consisting of the region that was inherited from the B.1.1.7 parental in the recombinant(s), and one consisting of the region that was inherited from the other parental. This resulted in 16 alignments in total. We reconstructed the phylogenetic relationships for each with IQTREE v2.1 (Minh et al. 2020), using the HKY model of nucleotide substitution, conducting 1000 ultrafast bootstrap replicates (Minh, Nguyen, and von Haeseler 2013; Hoang et al. 2018), and rooting the tree on the reference sequence, which is basal to all B lineage sequences. The phylogenetic trees produced by this analysis are available at https://github.com/COG-UK/UK-recombination-analysis.

To determine the placement of the different regions of each recombinant genome in a single context, we also built a phylogenetic tree of the representative background’s complete genomes, to which we added the masked recombinant genomes, so that each recombinant was present in the alignment twice, once with the B.1.1.7 region of its genome unmasked, and once with the opposing region unmasked. We ran IQTREE as above.

### Follow up of putative recombinants

To test for onward community transmission of the putative recombinants, we searched the whole UK dataset as of the 5th May 2021 for additional sequences whose genetic variation matched the variation of the recombinants. For each of the eight set of recombinants, we defined a set of SNPs and deletions by which all the recombinants within that set differed from the reference sequence (MN908947.3). Then we used type_variants to scan the UK dataset for genomes whose SNP and deletion variation was compatible with being a descendant or sibling of the putative recombinants. Group A represented the only recombination event with evidence for further transmission according to the results of this procedure. We carried out the following additional analyses to further investigate transmission of Group A genomes. Firstly, we visualised the nucleotide variation of the additional matching genomes using snipit and extracted their sampling locations and dates. Secondly, to explore the phylogenetic context of Group A and its derivatives, we reconstructed their (whole-genome) phylogenetic relationships using IQTREE. We also extracted the 100 closest sequences by genetic similarity for each alternate region of the genome (B.1.1.7-like and non-B.1.1.7-like) for each of the four original members of Group A to provide phylogenetic context to the parental sequences. This resulted in a dataset of 216 sequences in total when the two groups of neighbours were combined, and duplicates removed. We reconstructed their (whole-genome) phylogenetic relationships with the IQTREE, as above. We labelled the phylogenetic tree of recombinants and the phylogenetic tree of parental sequences with the sampling date in number of epidemiological weeks (epiweeks) since the first epiweek of 2020 to assess the temporal context of the recombination event and subsequent transmission. We carried out a second follow up on the 1st June using the same procedure as above.

