## Supplementary Information for "Generation and transmission of inter-lineage recombinants in the SARS-CoV-2 pandemic"

**Jackson et al**

**Supplementary Information**

#### Supplementary Figure Legends

***Figure S1***

The distribution of minor allele frequencies from the read data for the 16 putative recombinants (red bars) and 20 samples suspected of being sequenced mixtures, either due to co-infection or laboratory contamination (grey bars). For each recombinant, the minor allele frequency is the mean across all sites that differ by a nucleotide change from the reference (MN908947.3) in it or either of its putative parentals by genetic similarity. For the mixtures, the minor allele frequency is the mean across the sites that differ by a nucleotide change from the reference at genomic positions where mutations occur in B.1.1.7.

***Figure S2***

The nucleotide variation with respect to the reference sequence (MN908947.3; grey genome far bottom) for each of the recombinant genomes (middle coloured genomes in each panel) and their closest neighbours by genetic similarity among all UK sequences from the same time period, for the B.1.1.7-like and non-B.1.1.7-like regions of their genomes (top and bottom coloured genomes in each panel). Panels: A) Group B; B) Group C; C) Group D; D) CAMC-CBA018; E) CAMC-CB7AB3; F) MILK-103C712; G) QEUH-1067DEF.

***Figure S3***

The distribution of the most frequent SARS-CoV-2 lineages in the NUTS1 location of each set (Groups A-D and the four singletons) of recombinants for the four weeks immediately preceding each set's (earliest) sample date. Here, B.1.177 refers to B.1.177 itself and all its descendant lineages (e.g. B.1.177.9); the same is true for B.1.36.

***Figure S4***

The distribution of nucleotide variation in the original members of Group A (top four coloured rows) and the 41 additional sequences that are derived from it (bottom 41 coloured rows), with respect to the reference sequence (MN908947.3; very bottom grey sequence).

**Table S1**

Nucleotide, amino acid, or deletion states at the 22 sites where mutations define lineage B.1.1.7 from its immediate ancestor, for each putative recombinant. The genotype column names are in one of the following formats: (i) “snp”, followed by the nucleotide in MN908947.3 (reference), the position in the MN908947.3 genome, then the nucleotide in B.1.1.7; (ii) “aa” (amino acid), followed by the name of the coding sequence, the residue in MN908947.3, the position of the residue in the coding sequence, then the residue in B.1.1.7; (iii) “del” (deletion) followed by the first position of the deletion, then the length of the deletion. For the contents of the amino acid columns, an asterisk (*) denotes a stop codon. For the contents of the deletion genotype columns, del=deletion present; ref=deletion absent. Lineage B.1.1.7 states are coloured red, and MN908947.3 alleles are coloured white. The mosaic structures visible here informed the first iteration of recombination detection.

| Sample Name | Group | Sample Date | snp:C913T | aa:ORF1ab:T1001I | aa:ORF1ab:A1708D | snp:C5986T | aa:ORF1ab:I2230T | del:11288:9 | snp:C14676T | snp:C15279T | snp:T16176C | del:21765:6 | del:21991:3 | aa:s:N501Y | aa:s:A570D | aa:s:P681H | aa:s:T716I | aa:s:S982A | aa:s:D1118H | aa:ORF8:Q27* | aa:ORF8:R52I | aa:ORF8:Y73C | aa:N:D3L | aa:N:S235F |
|---|---|---|---|---|---|---|---|---|---|---|---|---|---|---|---|---|---|---|---|---|---|---|---|---|
| CAMC-CBA018 | - | 18/12/2020 | C | T | A | C | I | ref | C | C | T | del | del | Y | D | H | I | A | H | * | I | C | L | F |
| ALDP-11CF93B | A | 30/01/2021 | C | T | A | C | I | ref | C | C | T | del | del | Y | D | H | I | A | H | * | I | C | L | F |
| ALDP-125C4D7 | A | 06/02/2021 | C | T | A | C | I | ref | C | C | T | del | del | Y | D | H | I | A | H | * | I | C | L | F |
| LIVE-DFCFFE | A | 14/02/2021 | C | T | A | C | I | ref | C | C | T | del | del | Y | D | H | I | A | H | * | I | C | L | F |
| ALDP-130BB95 | A | 21/02/2021 | C | T | A | C | I | ref | C | C | T | del | del | Y | D | H | I | A | H | * | I | C | L | F |
| QEUH-CCCB30 | B | 23/12/2020 | C | T | A | C | T | del | T | T | C | del | del | Y | D | H | I | A | H | * | I | C | L | F |
| QEUH-CD0F1F | B | 24/12/2020 | C | T | A | C | T | del | T | T | C | del | del | Y | D | H | I | A | H | * | I | C | L | F |
| MILK-1166F52 | C | 24/01/2021 | T | I | D | T | T | del | T | T | C | del | del | Y | D | H | I | A | H | Q | R | Y | D | S |
| MILK-11C95A6 | C | 30/01/2021 | T | I | D | T | T | del | T | T | C | del | del | Y | D | H | I | A | H | Q | R | Y | D | S |
| QEUH-109B25C | C | 18/01/2021 | T | I | D | T | T | del | T | T | C | del | del | Y | D | H | I | A | H | Q | R | Y | D | S |
| MILK-126FE1F | D | 07/02/2021 | C | T | A | C | I | ref | C | C | T | ref | ref | Y | D | H | I | A | H | * | I | C | L | F |
| RAND-12671E1 | D | 02/02/2021 | C | T | A | C | I | ref | C | C | T | ref | ref | Y | D | H | I | A | H | * | I | C | L | F |
| RAND-128FA33 | D | 02/02/2021 | C | T | A | C | I | ref | C | C | T | ref | ref | Y | D | H | I | A | H | * | I | C | L | F |
| CAMC-CB7AB3 | - | 18/12/2020 | T | I | A | C | I | ref | C | C | T | del | del | Y | D | H | I | A | H | * | I | C | L | F |
| MILK-103C712 | - | 12/01/2021 | C | T | A | C | I | ref | C | C | T | ref | ref | N | A | P | T | S | D | * | I | C | L | F |
| QEUH-1067DEF | - | 17/01/2021 | T | I | D | T | T | ref | C | C | T | ref | ref | N | A | P | T | S | D | Q | R | Y | D | S |

**Table S2**

GISAID and ENA accession numbers for the 16 putative recombinant sequences.

| Sample | Group | GISAID accession | ENA sample accession | ENA sample secondary accession | ENA run accession |
|---|---|---|---|---|---|
| ALDP-11CF93B | A | EPI_ISL_989697 | ERS5764793 | SAMEA8077663 | ERR5308556 |
| ALDP-125C4D7 | A | EPI_ISL_1019487 | ERS5792357 | SAMEA8105282 | ERR5323237 |
| ALDP-130BB95 | A | EPI_ISL_1122630 | ERS5883487 | SAMEA8196877 | ERR5414941 |
| LIVE-DFCFFE | A | EPI_ISL_1104468 | ERS5872630 | | ERR5404883 |
| QEUH-CCCB30 | B | EPI_ISL_782203 | ERS5522602 | SAMEA7775362 | ERR5058070 |
| QEUH-CD0F1F | B | EPI_ISL_782442 | ERS5522673 | SAMEA7775433 | ERR5058141 |
| MILK-1166F52 | C | EPI_ISL_938901 | ERS5697106 | SAMEA8009867 | ERR5272107 |
| MILK-11C95A6 | C | EPI_ISL_1050799 | ERS5874115 | | ERR5406307 |
| QEUH-109B25C | C | EPI_ISL_909004 | ERS5669824 | SAMEA7982538 | ERR5232711 |
| MILK-126FE1F | D | EPI_ISL_1057512 | ERS5812369 | SAMEA8125397 | ERR5349458 |
| RAND-12671E1 | D | EPI_ISL_1045772 | ERS5805519 | SAMEA8118538 | ERR5335088 |
| RAND-128FA33 | D | EPI_ISL_1042110 | ERS5809401 | SAMEA8122425 | ERR5340986 |
| CAMC-CBA018 | - | EPI_ISL_777766 | ERS5517714 | SAMEA7770468 | ERR5054123 |
| CAMC-CB7AB3 | - | EPI_ISL_777974 | ERS5517673 | SAMEA7770427 | ERR5054082 |
| MILK-103C712 | - | EPI_ISL_994038 | ERS5760589 | SAMEA8073454 | ERR5304348 |
| QEUH-1067DEF | - | EPI_ISL_917983 | ERS5674718 | SAMEA7987439 | ERR5238288 |

**Table S3**

Phylogenetic placement and UF bootstrap support for the two parental tracts of each recombinant's genome, among the whole UK epidemic. For CAMC-CB7AB3, "Left" refers to the inner part of its genome, and "Right" refers to the outer part of its genome. The full phylogenies with bootstrap support are available at https://github.com/COG-UK/UK-recombination-analysis

| Sample | Group | Left placement | Left support | Right placement | Right support |
|---|---|---|---|---|---|
| ALDP-11CF93B | A | B.1.177 | 81 | B.1.1.7 | 100 |
| ALDP-125C4D7 | A | B.1.177 | 81 | B.1.1.7 | 100 |
| LIVE-DFCFFE | A | B.1.177 | 81 | B.1.1.7 | 100 |
| ALDP-130BB95 | A | B.1.177 | 81 | B.1.1.7 | 100 |
| QEUH-CCCB30 | B | B.1.36.28 | 97 | B.1.1.7 | 100 |
| QEUH-CD0F1F | B | B.1.36.28 | 97 | B.1.1.7 | 100 |
| MILK-1166F52 | C | B.1.1.7 | 100 | B.1.221.1 | 98 |
| MILK-11C95A6 | C | B.1.1.7 | 100 | B.1.221.1 | 98 |
| QEUH-109B25C | C | B.1.1.7 | 100 | B.1.221.1 | 98 |
| MILK-126FE1F | D | B.1.36.17/B.1.36.39 | 100 | B.1.1.7 | 100 |
| RAND-12671E1 | D | B.1.36.17/B.1.36.39 | 100 | B.1.1.7 | 100 |
| RAND-128FA33 | D | B.1.36.17/B.1.36.39 | 100 | B.1.1.7 | 100 |
| CAMC-CB7AB3 | - | B.1.177 | 39 | B.1.1.7 | 100 |
| CAMC-CBA018 | - | B.1.177 | 94 | B.1.1.7 | 100 |
| MILK-103C712 | - | B.1.177 | 100 | B.1.1.7 | 100 |
| QEUH-1067DEF | - | B.1.1.7 | 100 | B.1.177 | 100 |

**Figure S1**

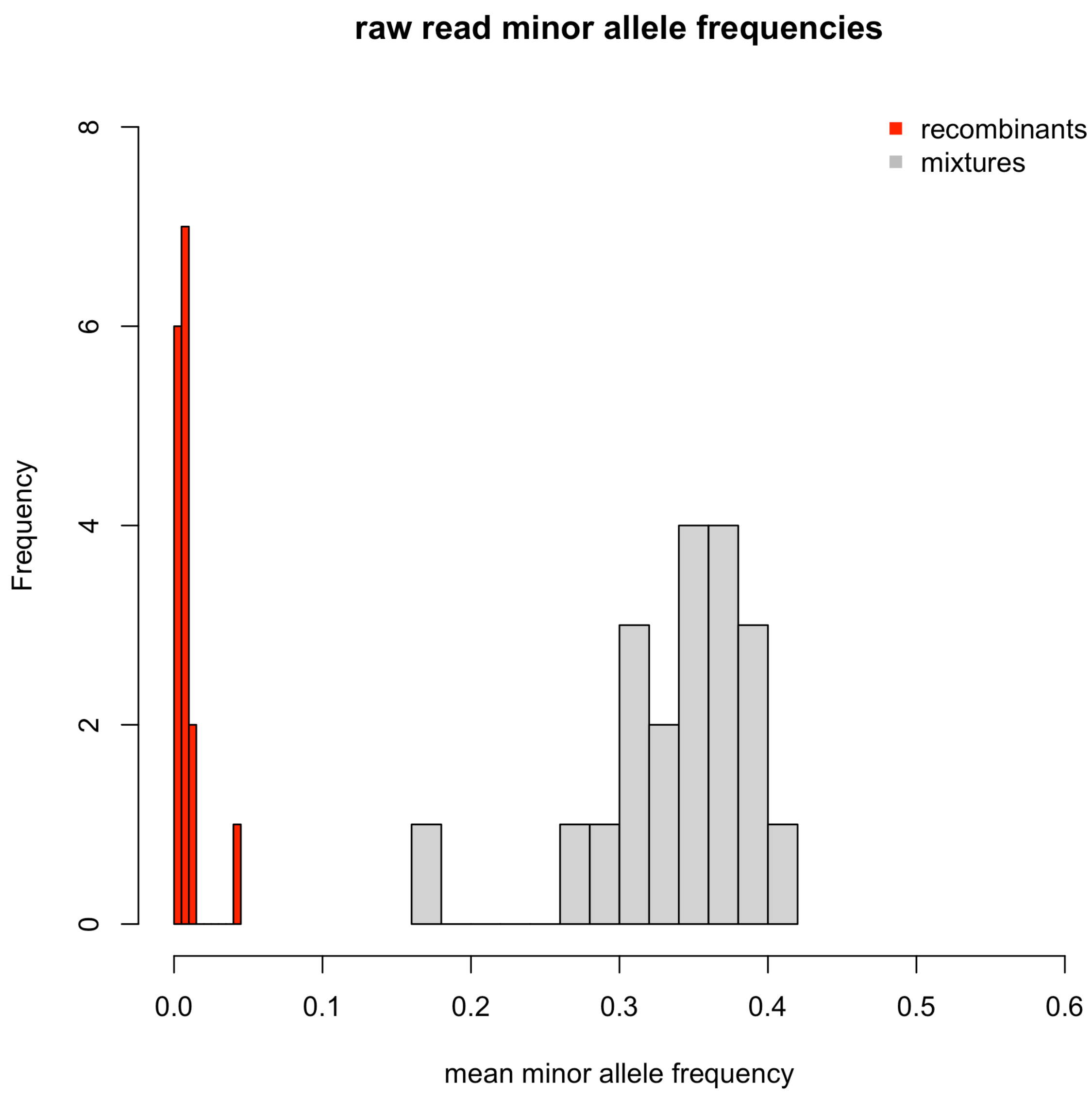
raw read minor allele frequencies
recombinants
mixtures
Frequency
0
2
4
6
8
0.0
0.1
0.2
0.3
0.4
0.5
0.6
mean minor allele frequency

### Figure S2

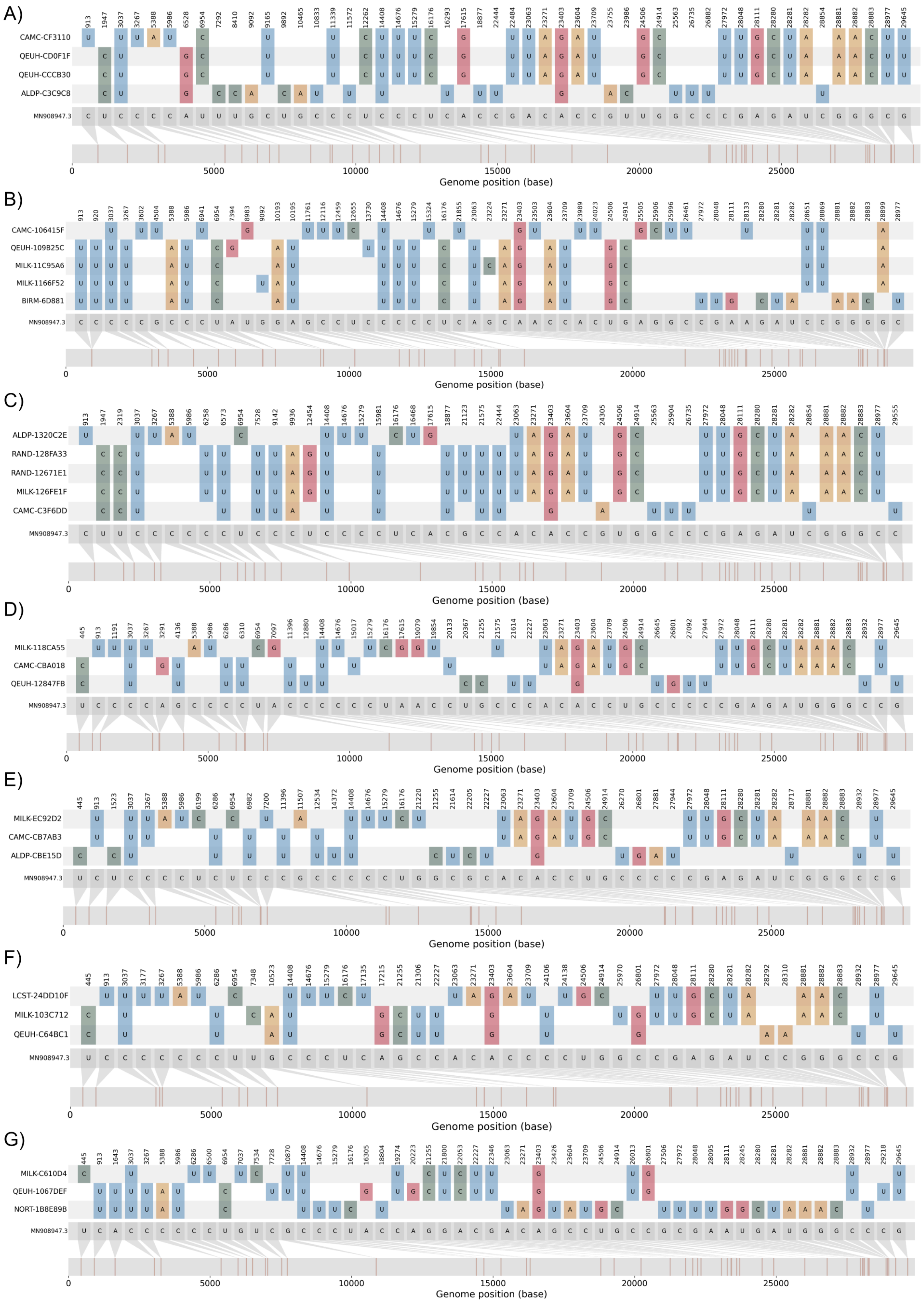

A)
CAMC-CF3110
QEUH-CD0F1F
QEUH-CCCB30
ALDP-C3C9C8
MN908947.3
Genome position (base)
B)
CAMC-106415F
QEUH-109B25C
MILK-11C95A6
MILK-1166F52
BIRM-6D881
MN908947.3
Genome position (base)
C)
ALDP-1320C2E
RAND-128FA33
RAND-12671E1
MILK-126FE1F
CAMC-C3F6DD
MN908947.3
Genome position (base)
D)
MILK-118CA55
CAMC-CBA018
QEUH-12847FB
MN908947.3
Genome position (base)
E)
MILK-EC92D2
CAMC-CB7AB3
ALDP-CBE15D
MN908947.3
Genome position (base)
F)
LCST-24DD10F
MILK-103C712
QEUH-C64BC1
MN908947.3
Genome position (base)
G)
MILK-C610D4
QEUH-1067DEF
NORT-1B8E89B
MN908947.3
Genome position (base)

**Figure S3**

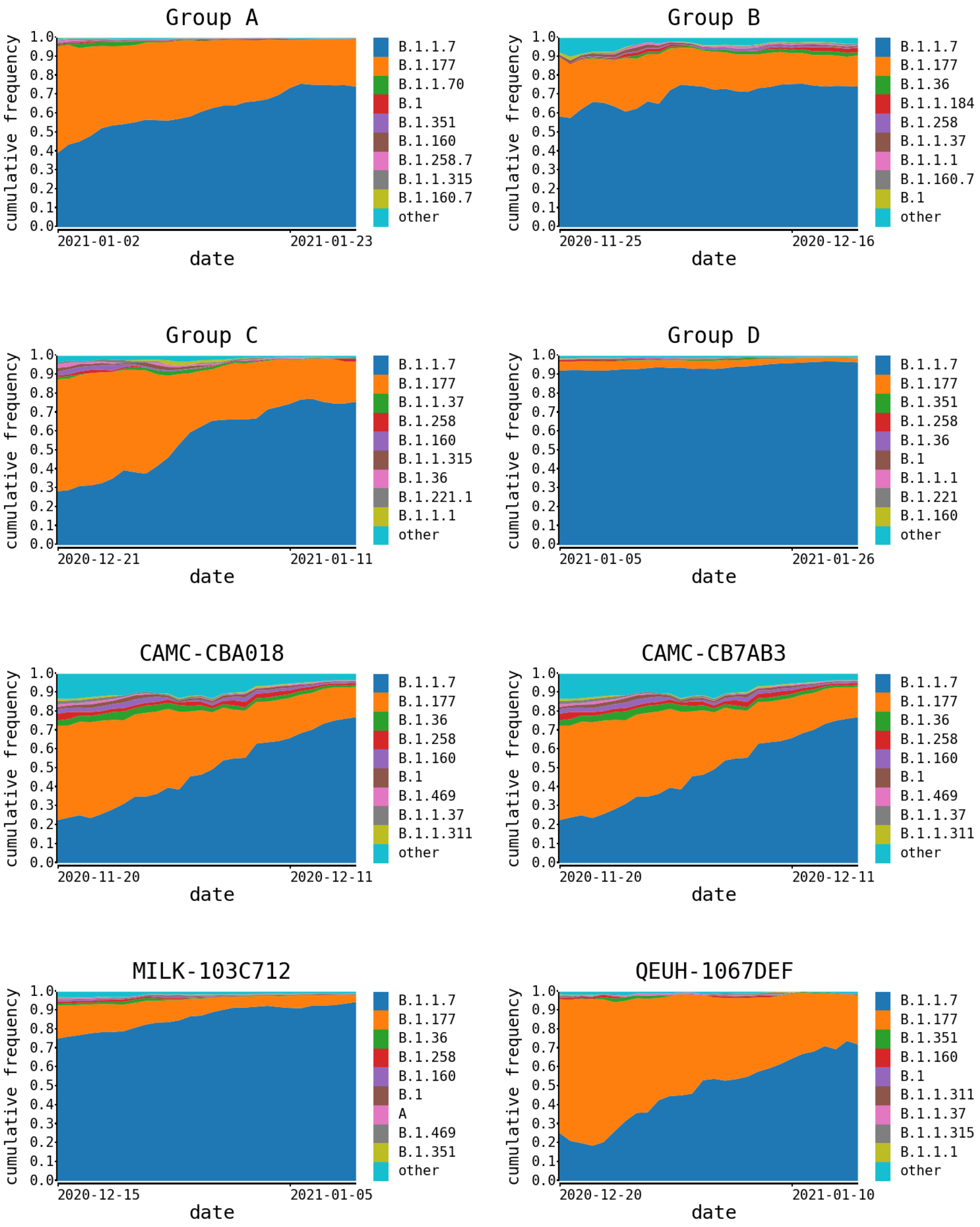

Group A
cumulative frequency
1.0
0.9
0.8
0.7
0.6
0.5
0.4
0.3
0.2
0.1
0.0
2021-01-02
2021-01-23
date
B.1.1.7
B.1.177
B.1.1.70
B.1
B.1.351
B.1.160
B.1.258.7
B.1.1.315
B.1.160.7
other
Group B
cumulative frequency
2020-11-25
2020-12-16
date
B.1.1.7
B.1.177
B.1.36
B.1.1.184
B.1.258
B.1.1.37
B.1.1.1
B.1.160.7
B.1
other
Group C
cumulative frequency
2020-12-21
2021-01-11
date
B.1.1.7
B.1.177
B.1.1.37
B.1.258
B.1.160
B.1.1.315
B.1.36
B.1.221.1
B.1.1.1
other
Group D
cumulative frequency
2021-01-05
2021-01-26
date
B.1.1.7
B.1.177
B.1.351
B.1.258
B.1.36
B.1
B.1.1.1
B.1.221
B.1.160
other
CAMC-CBA018
cumulative frequency
2020-11-20
2020-12-11
date
B.1.1.7
B.1.177
B.1.36
B.1.258
B.1.160
B.1
B.1.469
B.1.1.37
B.1.1.311
other
CAMC-CB7AB3
cumulative frequency
2020-11-20
2020-12-11
date
B.1.1.7
B.1.177
B.1.36
B.1.258
B.1.160
B.1
B.1.469
B.1.1.37
B.1.1.311
other
MILK-103C712
cumulative frequency
2020-12-15
2021-01-05
date
B.1.1.7
B.1.177
B.1.36
B.1.258
B.1.160
B.1
A
B.1.469
B.1.351
other
QEUH-1067DEF
cumulative frequency
2020-12-20
2021-01-10
date
B.1.1.7
B.1.177
B.1.351
B.1.160
B.1
B.1.1.311
B.1.1.37
B.1.1.315
B.1.1.1
other

**Figure S4**

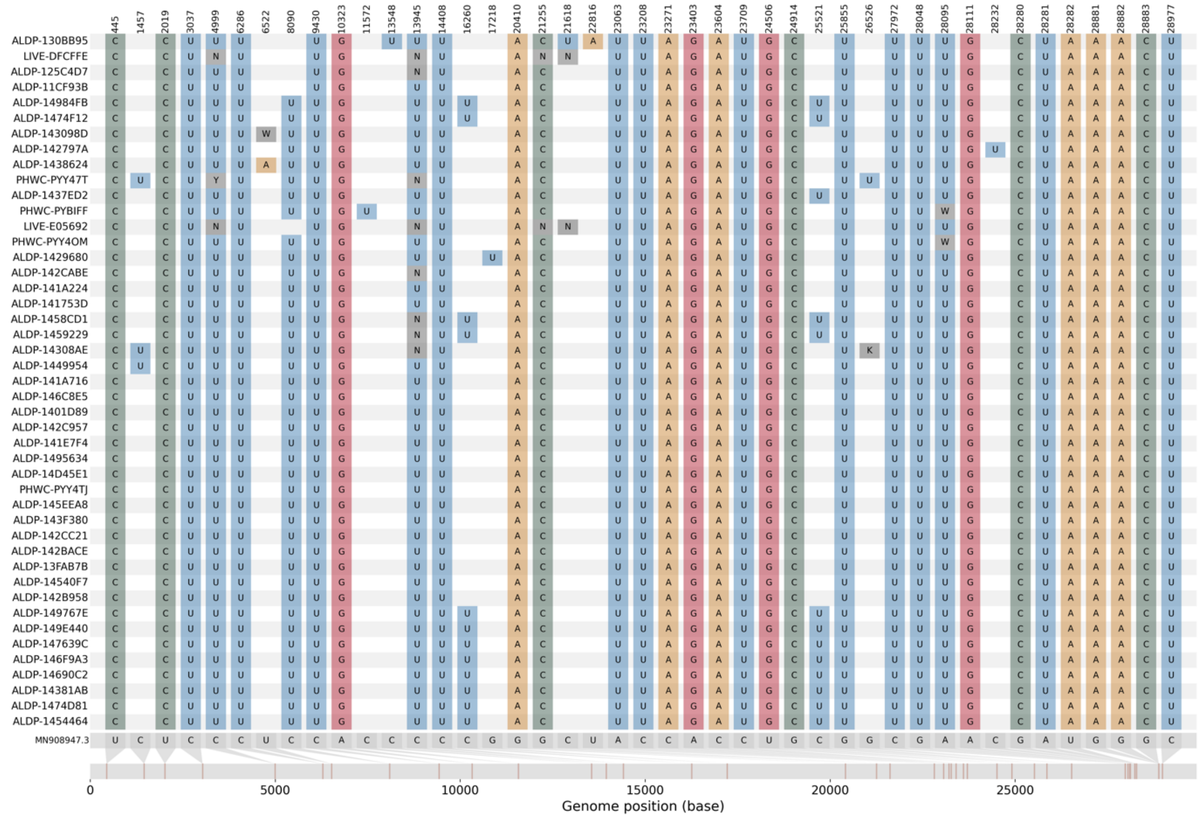
445
1457
2019
3037
4999
6286
6522
8090
9430
10323
11572
13548
13945
14408
16260
17218
20410
21255
21618
22816
23063
23208
23271
23403
23604
23709
24506
24914
25521
25855
26526
27972
28048
28095
28111
28232
28280
28281
28282
28881
28882
28883
28977
ALDP-130BB95
LIVE-DFCFFE
ALDP-125C4D7
ALDP-11CF93B
ALDP-14984FB
ALDP-1474F12
ALDP-143098D
ALDP-142797A
ALDP-1438624
PHWC-PYY47T
ALDP-1437ED2
PHWC-PYBIFF
LIVE-E05692
PHWC-PYY4OM
ALDP-1429680
ALDP-142CABE
ALDP-141A224
ALDP-141753D
ALDP-1458CD1
ALDP-1459229
ALDP-14308AE
ALDP-1449954
ALDP-141A716
ALDP-146C8E5
ALDP-1401D89
ALDP-142C957
ALDP-141E7F4
ALDP-1495634
ALDP-14D45E1
PHWC-PYY4TJ
ALDP-145EEA8
ALDP-143F380
ALDP-142CC21
ALDP-142BACE
ALDP-13FAB7B
ALDP-14540F7
ALDP-142B958
ALDP-149767E
ALDP-149E440
ALDP-147639C
ALDP-146F9A3
ALDP-14690C2
ALDP-14381AB
ALDP-1474D81
ALDP-1454464
MN908947.3
0
5000
10000
15000
20000
25000
Genome position (base)
